# The impact of shielding during the COVID-19 pandemic on mental health: Evidence from the English Longitudinal Study of Ageing

**DOI:** 10.1101/2021.12.16.21267914

**Authors:** Giorgio Di Gessa, Debora Price

## Abstract

**Background:** During the COVID-19 pandemic, older and clinically vulnerable people were instructed to shield or stay at home to save lives. Policies restricting social contact and human interaction pose a risk to mental health, but we know very little about the impact of shielding and stay at home orders on the mental health of older people.

**Aims:** Understand the extent to which shielding contributes to poorer mental health.

**Method:** Exploiting longitudinal data from Wave 9 (2018/19) and two COVID-19 sub-studies (June/July 2020; November/December 2020) of the English Longitudinal Study of Ageing we use logistic and linear regression models to investigate associations between patterns of shielding during the pandemic and mental health, controlling for socio-demographic characteristics, pre-pandemic physical and mental health, and social isolation measures.

**Results:** By December 2020, 70% of older people were still shielding or staying at home, with 5% shielding throughout the first 9 months of the pandemic. Respondents who shielded experienced worse mental health. Although prior characteristics and lack of social interactions explain some of this association, even controlling for all covariates, those shielding throughout had higher odds of reporting elevated depressive symptoms (OR=1.87, 95%CI=1.22;2.87) and reported lower quality of life (B=-1.28, 95%CI=-2.04;-0.52) than those who neither shielded nor stayed at home. Shielding was also associated with increased anxiety.

**Conclusions:** Shielding itself seems associated with worse mental health among older people, highlighting the need for policymakers to address the mental health needs of those who shielded, both in emerging from the current pandemic and for the future.

## 1. Introduction

Early in the COVID-19 pandemic we knew that risks of serious illness and death increased exponentially with age, and that many diseases, also strongly correlated with age, increased morbidity and mortality risks [1]. When the UK government announced the first lockdown, shielding and stay-at-home orders were issued for at-risk groups. Some 3.8 million people in the UK were ordered to shield (almost 6 per cent of the population), 74 per cent of whom are over 50 [2]. Furthermore, from March 2020, all those over 70 *and* those deemed ‘clinically vulnerable’ were also advised to stay indoors and limit their interactions with others for 12 weeks. Restrictions were partially eased for a few months in the second half of 2020 but a new lockdown in England was reintroduced in November 2020 for a month, with a third in January 2021. Although shielding advice ended on 1^st^ April 2021, shielders were nevertheless recommended to continue to take precautions. Updated government Guidance published on 28^th^ July 2021 warned this group that vaccines are not 100% effective, may not work well in the immune-compromised, cautioned those at higher risk of becoming seriously ill if they were to catch COVID-19 and advised them to think carefully about contacts with other people [3]. Many people have avoided social contact throughout the pandemic regardless of formal advice [4, 5] and as variants circulate, many of those previously warned of their vulnerabilities to COVID-19 may elect to continue to protect themselves from social contact [6].

Policies restricting social contact and human interaction have clearly posed a risk to mental health and wellbeing [6-10], and research has demonstrated deteriorating mental health especially among those with pre-existing mental or physical health conditions and low social support [11-15]. Because the shielding policies and stay-at-home advice disproportionately impacted older people, researchers have also investigated mental health sequelae of the pandemic for this group. Despite some evidence of coping strategies [16], there has been a substantial deterioration in mental health and wellbeing for the over 50s during the pandemic [17-21], which has been shown to have been exacerbated by shielding for those identified as clinically vulnerable to COVID-19 [22, 23].

However, important lacunae remain in our understanding of poorer mental health in older people instructed to shield during the pandemic. First, not all people advised to shield did so [4, 23], and similarly individuals might have decided to shield even without such advice, particularly if they self-identified as at increased risk of serious illness or death [6]. It is therefore important to consider people’s behaviours and their interaction with mental health to better understand the role of shielding. Second, to our knowledge, this is the first study that considers reported behaviours at three time points that cover the first 8/9 months of the pandemic (two of which were characterised by ongoing lockdowns). Third, people advised to shield are also disproportionately likely to have had poorer physical, mental and social wellbeing prior to the pandemic. This study therefore also accounts for pre-pandemic characteristics which could potentially lead to a higher risk of social isolation and care deficits and therefore to deteriorating mental health and wellbeing. Finally, as one possible consequence of both shielding and staying at home is for physical separation to lead to social isolation and loneliness, this study also accounts for these important factors to understand the impact of shielding on mental health.

Identifying the effect of shielding on mental health is key to devising appropriate and targeted policy responses as we aim to build back society and restore the wellbeing of our populations, and in the event of the need for further lockdowns or future pandemics. Therefore, in this paper, we aim to understand the extent to which shielding and staying at home are *additional* factors contributing to poorer mental health above and beyond more traditional explanations such as socio-economic and demographic factors, prior health, or increased isolation and lack of social support during the pandemic.

## 2. Methods

### 2.1 Study Population

We used the most recent pre-pandemic data (wave 9, collected in 2018/19) and the two waves of the COVID-19 sub-study (collected in June/July and November/December 2020 respectively) of the English Longitudinal Study of Ageing (ELSA)[24]. ELSA is a longitudinal biennial survey representative of individuals aged 50 and over in private households. During the pandemic, 9,392 ELSA members were invited to participate online or by CATI (Computer-Assisted Telephone Interviewing) to the COVID-19 sub-study (75% response rate in both waves, 94% longitudinal response rate). Analyses were based on core respondents who participated in both COVID-19 waves with available information in Wave 9 (N=5,146). Further details of the survey’s sampling frame and methodology can be found at www.elsa-project.ac.uk. ELSA was approved by the London Multicentre Research Ethics Committee (MREC/01/2/91), with the COVID-19 sub-study approved by the UCL REC. Informed consent was obtained from all participants. All data are available through the UK Data Service (SN 8688 and 5050).

### 2.2 Main measurements of interest

#### Shielding

Respondents were asked whether in April 2020 they were shielding (‘not leaving home for any reason, not going out to buy food and not seeing people outside of your household’); staying at home (‘only leaving your home for very limited purposes, such as shopping for food, one form of exercise, or essential work’); or neither. Similar questions were asked at both COVID-19 waves with respect to the week prior to the interview. Based on the possible combinations of answers, we classified respondents into 5 broad categories distinguishing between those who, during the first 8 – 9 months of the pandemic: (i) were staying at home (but not shielding) at all three measured time points; (ii) were shielding at all three time points; (iii) shielded in two time periods (and were mostly staying at home in the third); (iv) shielded in one time period (and described themselves as either staying at home or neither shielding nor staying at home in the other two); and (v) were not shielding at any point, and did not stay at home throughout the period.

#### Mental Health

We considered four outcome measures of mental health assessed at the second COVID-19 wave: depressive symptoms, anxiety, well-being and quality of life. Symptoms of depression were measured by an abbreviated version of the validated Centre for Epidemiologic Studies Depression (CES-D) Scale [25]. The CES-D scale is not a diagnostic instrument for clinical depression but can be used to identify people “at risk” of depression in population-based studies [26]. This short version has good internal consistency (Cronbach’s α >0.95) and comparable psychometric properties to the full 20-item CES-D [27]. The scale includes 8 binary (no/yes) questions that enquire about whether respondents experienced any depressive symptoms, such as feeling sad or having restless sleep, in the week prior to interview. We classified respondents who reported four or more depressive symptoms on the CES-D scale as with elevated depressive symptoms [28, 29]. Anxiety was monitored with the Generalised Anxiety Disorder assessment (GAD-7), which evaluates the presence in the past two weeks of seven symptoms of anxiety, such as becoming easily annoyed or irritable or not being able to stop or control worrying, on a 4-point scale (“Not at all, “Several days”, “More than half the days”, “Nearly every day”). This is a well-validated tool, with a high scale reliability (Cronbach α=0·90 in this study) used to screen for generalised anxiety disorder in clinical practice and research [30]. A standard threshold score of 10 on the GAD-7 scale was used to define clinically significant symptoms. Furthermore, we considered subjective quality of life (QoL) evaluated using the CASP-12 scale. This is an abbreviated measure of the validated CASP-19 scale which was specifically designed for individuals in later life and used in a wide variety of ageing surveys [31]. CASP-12 contains 12 Likert-scaled questions measuring older people’s control and autonomy as well as self-realization through pleasurable activities. The possible range of CASP-12 scores is from 0 to 36, with higher scores indicating greater well-being; CASP-19 is treated as a continuous variable. Finally, we considered life satisfaction as a measure of personal well-being assessed using the Office for National Statistics (ONS) well-being scale (“On a scale of 0 to 10, where 0 is “not at all” and 10 is “very”, how satisfied are you with your life nowadays?”). This allows respondents to integrate and weigh various life domains the way they choose [32].

#### Covariates

Our analyses controlled for a wide range of demographic, socio-economic characteristics, health, and social support characteristics. We controlled for age and age squared to account for non-linear relationships with the outcome variables; sex; and ethnicity (White vs non-White participants due to data constraints in ELSA). To capture respondents’ socio-economic characteristics we controlled for pre-pandemic education, income, wealth, housing tenure and paid employment during the pandemic. Educational level was recoded into low (below secondary), middle, and high (university or above) following the International Standard Classification of Education (http://www.uis.unesco.org/). We categorised respondents by quintiles of wealth (total net non-pension non-housing wealth) and accounted for their equivalised total income (from paid work, state benefits, pensions and assets). Housing tenure distinguished outright owners, owners with a mortgage, and non-owners. Paid employment distinguished retired, in paid work and not working from home, in paid work and mostly working from home, furloughed, and other (including homemakers, unemployed, and sick or disabled).

We also accounted for pre-pandemic health. In particular, we controlled for disability (having impairments with basic and instrumental activities of daily living) and clinical vulnerability to COVID-19 (defined irrespective of age as reporting chronic lung disease, asthma, coronary heart disease, Parkinson’s disease, multiple sclerosis, diabetes; weakened immune system as a result of cancer treatment in the previous two years; BMI of 40 or above; or having been advised to shield by their GP/NHS)[22, 33]. We further controlled for pre-pandemic measures of mental health (see above for derivation). For GAD-7 —not included in pre-pandemic waves —analyses were adjusted for pre-pandemic ratings on the ONS anxiety scale.

Finally, we included indicators of social isolation and social support during the pandemic including household composition; social contacts; and loneliness. For household composition, we distinguished between respondents living alone; with a partner only; with partner and child(ren); with child(ren) but not the partner; and any other arrangements. Both COVID-19 ELSA surveys asked questions about real-time contact (by telephone or video calling) with family outside the household and with friends in the past month. At each wave, we categorised respondents as having infrequent contact if they reported contact with family and friends less than once a week or never. We constructed a variable indicating if respondents never reported infrequent contact; only at one time point; or at both COVID-19 waves. Finally, using the short version of the Revised UCLA loneliness scale with scores of 6 and higher indicating greater loneliness [34], we created a variable indicating whether respondents never felt lonely during the pandemic; felt lonely only at one point; or reported significant loneliness at both COVID-19 waves.

### 2.3 Statistical analysis

Following descriptive analysis, we investigated the longitudinal associations between shielding and mental health using nested logistic or linear models depending on the outcome. Following a ‘basic’ adjustment model (Model 1) which controlled for age, age squared, sex, and ethnicity, three further models were performed, with each including variables from the previous model. Model 2 adjusted for socio-economic characteristics (education, income, wealth, home tenure, and employment) as well as for pre-pandemic health (disability, and clinical vulnerability), as health conditions might have triggered the decision to shield or stay at home throughout the pandemic. Model 3 further adjusted for pre-pandemic relevant mental health measures. Finally, in Model 4, we adjusted for household composition, social contacts, and loneliness to explore whether and to what extent the relationships observed between shielding and mental health may be driven by reduced social interactions and higher loneliness during the pandemic. All analyses were performed using Stata 16. Cross-sectional and longitudinal sampling weights were employed to account for different probabilities of being included in the sample and for nonresponse to the survey.

## 3. Results

### 3.1 Descriptive statistics

The characteristics of the respondents are shown in Supplementary Table S1. Figure 1 shows that overall, the percentage of ELSA respondents who reported ‘shielding’ during the 2020 pandemic declined over time, with 22% of the sample shielding in April compared to 15% in June/July and 12% in November/December. Similarly, the percentage of the sample who left their homes only for limited purposes decreased from 63% in April to 57% in November/December 2020. Those who were doing neither increased from 15% in April 2020 to 28% by November/December. Table 1 shows the patterns of shielding behaviour over time. About 28% of respondents reported that they shielded at least once, with 5% shielding throughout; 11% on two (mostly consecutive) occasions; and 12% only once (mostly early in the pandemic). Among those who never reported shielding, respondents were equally split between those who were staying at home all the time (35%) and those who did not stay at home throughout the period (37%). A more comprehensive table with more detailed patterns of shielding/staying at home behaviours can be found in Supplementary Table S2.

**Figure 1.**
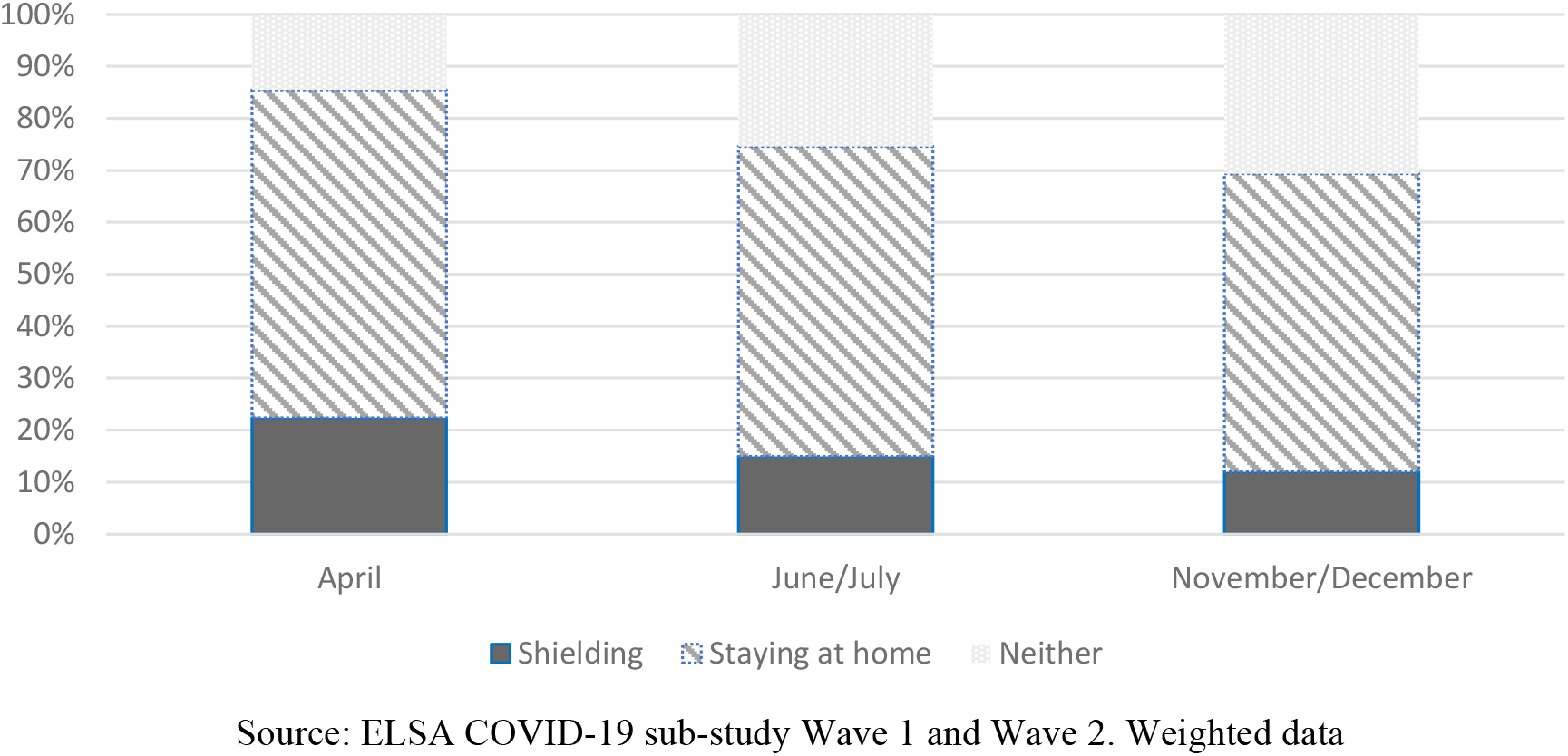
Percentages shielding, staying at home or neither in each ELSA Wave.

**Table 1.**
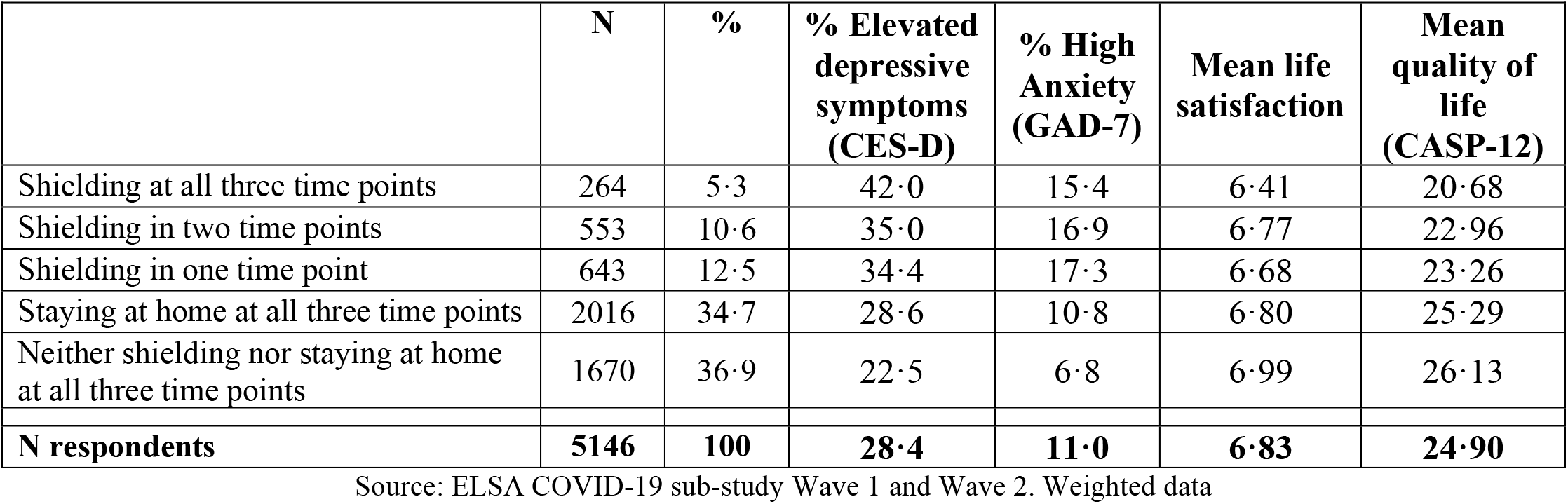
Distribution of patterns of shielding and staying at home across three time points (April 2020, June/July 2020 and November/December 2020) and Unadjusted mental health by patterns of shielding and staying at home.

|  | N | % | % Elevated depressive symptoms (CES-D) | % High Anxiety (GAD-7) | Mean life satisfaction | Mean quality of life (CASP-12) |
| --- | --- | --- | --- | --- | --- | --- |
| Shielding at all three time points | 264 | 5.3 | 42.0 | 15.4 | 6.41 | 20.68 |
| Shielding in two time points | 553 | 10.6 | 35.0 | 16.9 | 6.77 | 22.96 |
| Shielding in one time point | 643 | 12.5 | 34.4 | 17.3 | 6.68 | 23.26 |
| Staying at home at all three time points | 2016 | 34.7 | 28.6 | 10.8 | 6.80 | 25.29 |
| Neither shielding nor staying at home at all three time points | 1670 | 36.9 | 22.5 | 6.8 | 6.99 | 26.13 |
| **N respondents** | **5146** | **100** | **28.4** | **11.0** | **6.83** | **24.90** |

Source: ELSA COVID-19 sub-study Wave 1 and Wave 2. Weighted data

Table 1 also shows that mental health measured in the second wave of the COVID-19 ELSA sub-study showed substantial variation by shielding patterns. Respondents who shielded at all times reported the highest percentages of elevated depressive symptoms (42%), the lowest mean life satisfaction (6.4), and the lowest quality of life (Mean CASP-12=20.7), whereas those who neither shielded nor stayed at home throughout the period reported the best mental health and wellbeing (with 23% reporting elevated depressive symptom, 7% Anxiety; mean life satisfaction 7; and mean CASP-12 quality of life 26.1).

### 3.2 Multivariable analyses

To investigate how patterns of shielding and staying at home during the pandemic were associated with mental health we used multiple logistic (Table 2) and linear regressions (Table 3) and present results for the main variable of interest from four nested models (Full results in Supplementary Tables S3-S6). Accounting for basic demographic characteristics (Model 1), there were significant differences in mental health by patterns of self-isolation and shielding. Participants who shielded had higher odds of elevated depressive symptoms and anxiety and lower levels of quality of life and life satisfaction, with those shielding throughout reporting poorer mental health outcomes. Respondents who left their home only for essential reasons throughout were also more likely to report poorer mental health on all outcomes than those who did not have this restriction.

**Table 2.**
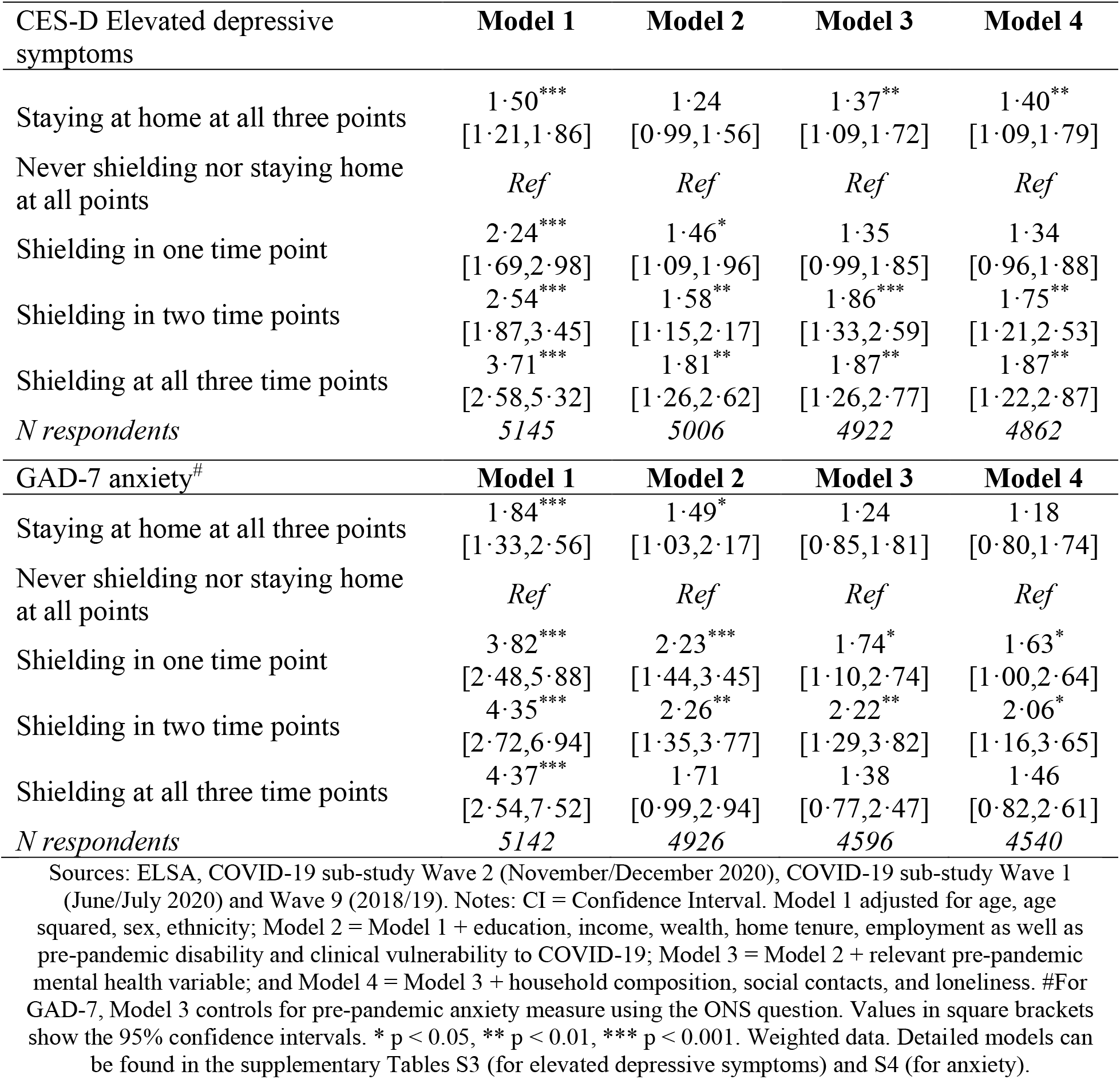
Associations between shielding patterns and elevated depressive symptoms and anxiety. Nested fully-adjusted logistic regression models – Odds Ratios [and 95% CIs].

**Table 3.**
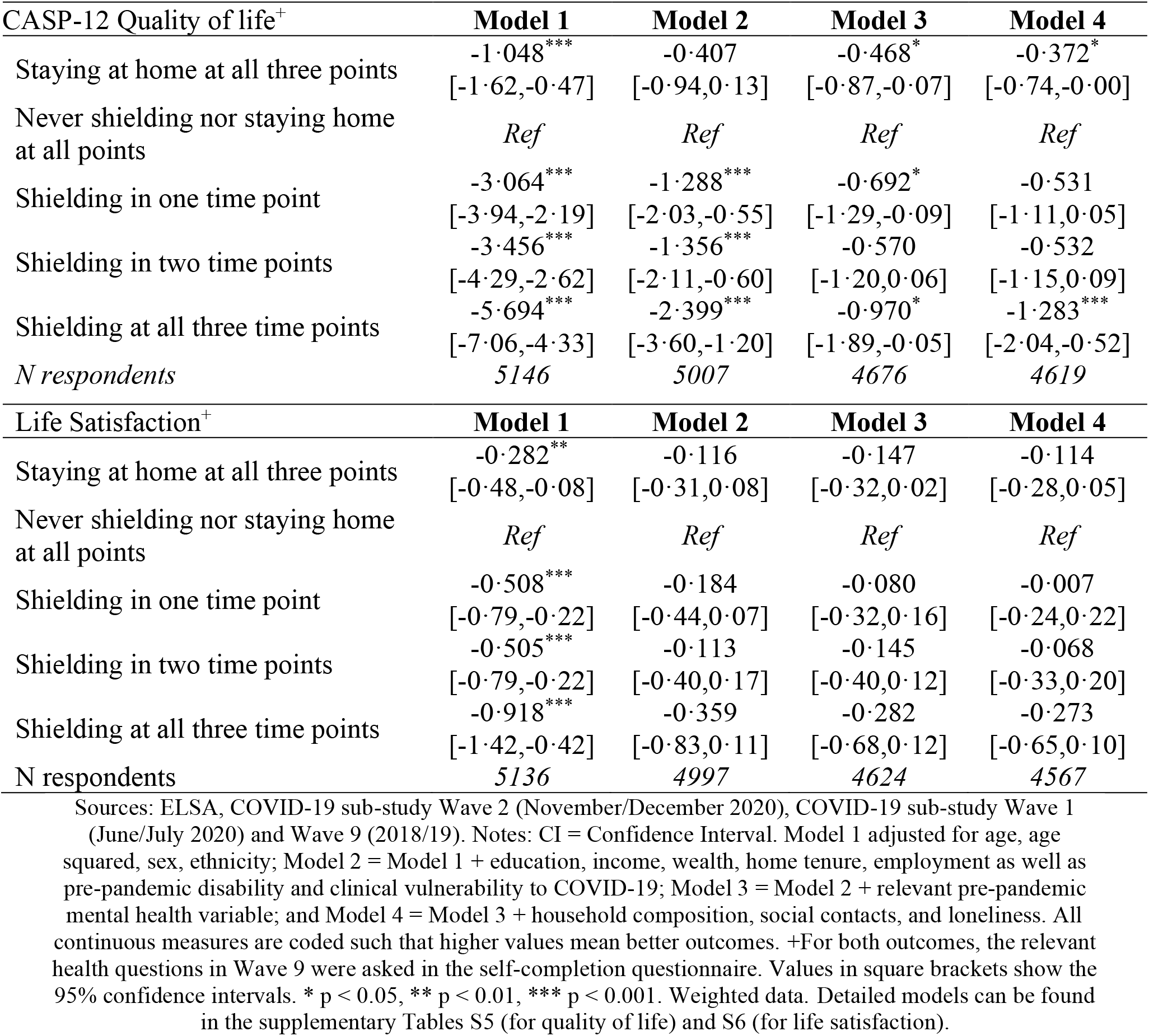
Associations between shielding patterns and quality of life and life satisfaction. Nested fully-adjusted linear regression models – Beta coefficients [and 95% CIs]

Results from Model 2, which additionally accounts for economic characteristics and pre-pandemic physical health, show that associations between patterns of shielding and poorer mental health were attenuated but remained significant for depressive symptoms, anxiety, and quality of life. However, all associations between patterns of shielding and life satisfaction are largely explained by respondents’ economic and pre-pandemic physical health. Moreover, accounting for these characteristics explained the association between shielding throughout the pandemic and anxiety.

In Model 3 we find that further adjusting for prior mental health, to account for the fact that those shielding at any time were more likely to have started the pandemic with worse mental health, made little difference to the observed associations between patterns of shielding, depressive symptoms, anxiety, and quality of life. For instance, those shielding at all three time points had higher odds of reporting depressive symptoms (OR=1.87, 95%CI=1.26-2.77) and reported lower quality of life (B=-0.97, p<0.05). These results remain largely robust to the addition of household composition, social contacts, and loneliness (Model 4), suggesting that little to none of the explanation for the relationship between shielding and mental health and wellbeing outcomes is explained by those shielding being lonelier or having reduced social contacts.

## 4. Conclusions and discussion

It is very important to understand the impact that shielding and stay at home policies, and the behavioural responses to them, have had on the mental health and wellbeing of different segments of the population. This is so not only for this pandemic but for the management of future outbreaks. In this paper, using data which gathered information about shielding, staying at home, or neither at three time points in 2020 (April, June/July, and November/December), we show that although over time fewer people aged 50 and older shielded or stayed at home, in November/December 2020 more than 70% of them described themselves as doing one or the other. Overall, staying at home throughout the pandemic or shielding at all (whether at one point, two or all three) were strongly associated with greater risk of elevated depressive symptoms, anxiety, poorer quality of life and lower life satisfaction. We also found an overall dose-response relationship between the frequency of shielding and poorer mental health, with those shielding at all times reporting the worse mental health outcomes once basic demographic characteristics were accounted for.

By using nested models we examined a number of hypothesised causes for this association – those shielding or staying at home coming disproportionately from more disadvantaged socio-economic positions, having a greater likelihood of having specific clinical conditions and living with disabilities, having poorer prior mental health, or having poorer social connections during the pandemic. Our models support this set of explanations to a limited degree. Socio-economic position and physical health do indeed substantially attenuate the relationships between shielding and poorer mental health, and provide the whole of the explanation for lower life satisfaction among those shielding or staying at home. Prior mental health also explains some of the association observed between shielding and poorer mental health. However, contrary to what might have been anticipated, social wellbeing indicators of loneliness, lack of social contact and household structure are not part of the explanatory matrix for this association and add little to the explanation. Even controlling for a number of possible mediating and confounding factors, strong and significant associations remain between staying at home or shielding at all times during the pandemic and higher depressive symptoms, and lower quality of life. For anxiety, however, we found heightened odds only for those who shielded once or twice during the pandemic, whereas the initial associations for those staying at home or shielding at all times were mostly explained by pre-pandemic anxiety. This provides support for the idea that the acts of shielding or staying at home themselves have a negative impact on mental health among older people as heralded by Webb [6] who, at the start of the pandemic, argued that the COVID-19 lockdown was “a perfect storm for older people’s mental health”. This study draws strength from using longitudinal data from the nationally-representative English Longitudinal Study of Ageing. It is the first to consider how the behavioural responses of older people to policy directives over time has contributed to mental health and wellbeing, and to test a variety of possible explanations for the association between shielding/staying at home and mental health outcomes. Our analysis supports the idea that shielding itself has been harmful, over and above other known vulnerabilities – perhaps because of the psychological impact of being told so starkly of your own vulnerability and mortality and the policing of your own behaviour, and associated anxiety and stress, that could plausibly result from that. Our contribution, however, should be considered in light of some limitations. ELSA did not collect information about respondents’ perception on their (lack of) independence during the pandemic, exposure to COVID-19 related news, individuals’ ability to tolerate and cope with the uncertainty due to COVID-19, or personality characteristics such as degree of risk tolerance or harm avoidance. These factors might help further understand both different behaviours and choices around levels of shielding and their subsequent effect on mental health. Also, although instructions to shield were mostly targeting older people, we could not evaluate associations across the full adult-age spectrum as ELSA samples only the over-50s, and those in care homes are excluded. Also, we only had information about shielding behaviours at three points in time, mostly referring to the week prior to the interview. While we cannot construct more nuanced and continuous measures of shielding/staying at home, this is likely to be the best data obtainable at scale for behaviours among older people during the pandemic. ELSA is limited to the population of England and so it is not possible to say that this would hold in other countries, although it is plausible that it would. Finally, ELSA suffers from non-random cumulative attrition, an unavoidable problem in longitudinal studies which can only partially be corrected for by using weights in the analysis.

In summary, our study provides a picture of the broader consequences of the pandemic and shielding policies among older people. While it is important to recognise that the clear aim and main benefit of guidelines focussing on social distancing is to contain the spread of the disease and save lives, policymakers need to be aware of adverse consequences for the mental health and well-being of those advised to shield or to stay at home. If the long-term health and social well-being of older people are not to be compromised by shielding and stay-at-home advice, urgent attention should be paid to addressing the mental health and wider needs of these groups, both in emerging from the current pandemic and if shielding policies remain a core strategy to protect individuals at higher risk from COVID-19 variants or indeed in a future pandemic.

## Data Availability

All data produced in the present study are available upon reasonable request to the authors

## Authors’ details

### Data Availability

The data used in this study are available from the UK Data Service with access codes SN 8688 and 5050. http://doi.org/10.5255/UKDA-SN-5050-20

## Acknowledgements

The data were made available through the UK Data Archive. ELSA was developed by a team of researchers based at University College London, NatCen Social Research, the Institute for Fiscal Studies, the University of Manchester and the University of East Anglia. The data were collected by NatCen Social Research. The funding is currently provided by the National Institute of Aging in the United States, and a consortium of UK government departments co-ordinated by the National Institute for Health Research. Funding has also been received by the Economic and Social Research Council. The developers and funders of ELSA and the Archive do not bear any responsibility for the analyses or interpretations presented here.

