## Supplementary Table for "The impact of shielding during the COVID-19 pandemic on mental health: Evidence from the English Longitudinal Study of Ageing"

### *Supplementary Table S1. Sample Characteristics*

| Sociodemographic and economic characteristics | % (N) |
|---|---|
| Mean Age during pandemic (SD) | 67.09 (10.46) |
| Female | 52.9 (2,931) |
| Non-White | 7.2 (189) |
| High Educational Qualification<br>Middle Educational Qualification<br>Low Educational Qualification | 20.8 (1,288)<br>49.4 (2,552)<br>29.9 (1,240) |
| Highest wealth quintile<br>2nd wealth quintile<br>3rd wealth quintile<br>4th wealth quintile<br>Lowest wealth quintile | 20.5 (1,259)<br>20.6 (1,238)<br>20.3 (1,095)<br>19.6 (846)<br>19.0 (639) |
| Retired<br>Employed (working not from home)<br>Employed (working mostly from home)<br>Furloughed<br>Sick/Disabled/Unemployed/Other | 54.0 (3,649)<br>19.5 (589)<br>13.6 (486)<br>3.5 (119)<br>9.5 (299) |
| Home owner (outright)<br>Home owner (with mortgage)<br>Home renter | 65.3 (4,011)<br>17.6 (571)<br>17.2 (563) |
| **Pre-pandemic health characteristics (at Wave 9)** | |
| 4 or more CES-D depressive symptoms | 12.7 (518) |
| With a score greater than 8 on the anxiety scale | 7.3 (297) |
| Mean CASP-12 (SD) | 26.21 (6.32) |
| Mean life satisfaction (SD) | 7.39 (2.23) |
| Clinically vulnerable to Covid-19 | 38.7 (1,945) |
| Disabled | 22.2 (1,059) |
| **Social contacts and well-being** | |
| Living Alone<br>Living with Partner/Spouse only<br>Living with Partner/Spouse and child(ren)<br>Living with child(ren) but no Partner/Spouse<br>Any other living arrangement | 24.8 (1,354)<br>45.6 (2,771)<br>18.7 (612)<br>6.7 (236)<br>4.2 (172) |
| Has never reported infrequent contacts during pandemic<br>Has reported infrequent contacts during pandemic once<br>Has reported infrequent contacts during pandemic twice | 57.0 (4,488)<br>8.8 (401)<br>4.1 (198) |
| Has never reported high UCLA loneliness during pandemic<br>Has reported high UCLA loneliness during pandemic once<br>Has reported high UCLA loneliness during pandemic twice | 67.9 (3,639)<br>16.4 (763)<br>15.7 (741) |
| **Total number of respondents (N)** | **5,146** |

Source: ELSA, Wave 9 (2018-2019), COVID-19 Substudy Wave 1 (June/July 2020) and COVID-19 sub-study Wave 2 (November/December 2020). Weighted data

### *Supplementary Table S2. Detailed patterns of shielding, staying at home, or neither at three time points during the pandemic*

| Classification | April 2020 | June/ July 2020 | November/ December 2020 | N | Unweighted Percentage |
|---|---|---|---|---|---|
| Shielding at all three time points | Shielding | Shielding | Shielding | 264 | 5.13 |
| Shielding in two time points | Shielding | Shielding | Staying home | 400 | 7.77 |
| | Shielding | Shielding | Neither | 60 | 1.17 |
| | Shielding | Neither | Shielding | 16 | 0.31 |
| | Shielding | Staying home | Shielding | 66 | 1.28 |
| | Staying home | Shielding | Shielding | 10 | 0.19 |
| | Neither | Shielding | Shielding | 1 | 0.02 |
| Shielding in one time point | Shielding | Staying home | Staying home | 262 | 5.09 |
| | Shielding | Staying home | Neither | 40 | 0.78 |
| | Shielding | Neither | Staying home | 72 | 1.40 |
| | Shielding | Neither | Neither | 27 | 0.52 |
| | Staying home | Shielding | Staying home | 34 | 0.66 |
| | Staying home | Shielding | Neither | 11 | 0.21 |
| | Staying home | Staying home | Shielding | 161 | 3.13 |
| | Staying home | Neither | Shielding | 16 | 0.31 |
| | Neither | Shielding | Staying home | 1 | 0.02 |
| | Neither | Neither | Shielding | 13 | 0.25 |
| | Neither | Staying home | Shielding | 6 | 0.12 |
| Staying at home at all three time points | Staying home | Staying home | Staying home | 2,016 | 39.18 |
| Neither shielding nor staying at home at all three time points | Staying home | Staying home | Neither | 647 | 12.57 |
| | Staying home | Neither | Staying home | 260 | 5.05 |
| | Staying home | Neither | Neither | 236 | 4.59 |
| | Neither | Staying home | Staying home | 37 | 0.72 |
| | Neither | Staying home | Neither | 16 | 0.31 |
| | Neither | Neither | Staying home | 157 | 3.05 |
| | Neither | Neither | Neither | 317 | 6.16 |

Source: ELSA, Wave 9 (2018-2019), COVID-19 Substudy Wave 1 (June/July 2020) and COVID-19 sub-study Wave 2 (November/December 2020). The first three column specify whether the respondents reported shielding, staying at home, or neither at each time point. Unweighted data

### *Supplementary Table S3. Associations between shielding patterns and depression. Nested fully-adjusted logistic regression models – Odds Ratios [and 95% CIs]*

| | **Model 1** | **Model 2** | **Model 3** | **Model 4** |
|---|---|---|---|---|
| Staying at home at all three points | 1.50*** [1.21,1.86] | 1.24 [0.99,1.56] | 1.37** [1.09,1.72] | 1.40** [1.09,1.79] |
| Never shielding nor staying home at all points | Ref | Ref | Ref | Ref |
| Shielding in one time point | 2.24*** [1.69,2.98] | 1.46* [1.09,1.96] | 1.35 [0.99,1.85] | 1.34 [0.96,1.88] |
| Shielding in two time points | 2.54*** [1.87,3.45] | 1.58** [1.15,2.17] | 1.86*** [1.33,2.59] | 1.75** [1.21,2.53] |
| Shielding at all three time points | 3.71*** [2.58,5.32] | 1.81** [1.26,2.62] | 1.87** [1.26,2.77] | 1.87** [1.22,2.87] |
| Female | 1.64*** [1.37,1.95] | 1.69*** [1.41,2.04] | 1.57*** [1.30,1.90] | 1.32** [1.07,1.62] |
| Age | 0.97*** [0.96,0.98] | 0.97*** [0.96,0.99] | 0.98** [0.96,0.99] | 0.98* [0.97,1.00] |
| Age squared | 1.00 [1.00,1.00] | 1.00 [1.00,1.00] | 1.00 [1.00,1.00] | 1.00 [1.00,1.00] |
| Non-White | 1.70* [1.09,2.64] | 1.35 [0.83,2.18] | 1.06 [0.67,1.68] | 0.92 [0.57,1.49] |
| Medium education | | 1.07 [0.84,1.37] | 1.16 [0.90,1.49] | 1.18 [0.91,1.54] |
| Low education | | 0.94 [0.71,1.25] | 0.93 [0.70,1.25] | 1.18 [0.88,1.60] |
| 2nd lowest quintile | | 0.84 [0.62,1.15] | 0.85 [0.62,1.17] | 0.84 [0.60,1.18] |
| 3rd wealth quintile | | 0.81 [0.60,1.09] | 0.85 [0.62,1.16] | 1.04 [0.73,1.47] |
| 4th wealth quintile | | 0.77 [0.56,1.05] | 0.81 [0.59,1.12] | 0.99 [0.69,1.42] |
| Highest wealth quintile | | 0.74 [0.52,1.05] | 0.81 [0.56,1.16] | 0.94 [0.63,1.41] |
| Income | | 0.90* [0.83,0.98] | 0.90* [0.82,0.99] | 0.92 [0.84,1.01] |
| Employed not Working From Home (WFH) | | 0.85 [0.59,1.22] | 0.88 [0.62,1.26] | 0.87 [0.59,1.28] |
| Employed mostly WFH | | 1.05 [0.72,1.52] | 1.21 [0.83,1.75] | 1.10 [0.74,1.64] |
| Furloughed | | 1.56 [0.88,2.75] | 1.69 [0.95,2.99] | 1.89* [1.04,3.43] |
| Other employment | | 2.31*** [1.58,3.38] | 2.19*** [1.48,3.23] | 1.94** [1.28,2.93] |
| Homeowner with mortgage | | 1.13 [0.84,1.52] | 1.19 [0.88,1.61] | 1.22 [0.88,1.68] |
| Rented accommodation | | 1.33* [1.00,1.76] | 1.26 [0.94,1.70] | 1.30 [0.94,1.80] |
| Clinically vulnerable to COVID-19 | | 1.19 [0.98,1.44] | 1.06 [0.87,1.28] | 0.98 [0.79,1.20] |
| Disability | | 2.40*** [1.97,2.94] | 1.81*** [1.45,2.25] | 1.76*** [1.40,2.22] |
| Pre-pandemic Depressed | | | 5.85*** [4.48,7.65] | 3.32*** [2.45,4.49] |
| Living Alone | | | | 1.06 [0.83,1.36] |
| Living with partner and children | | | | 1.24 [0.92,1.69] |
| Single parent | | | | 1.42 [0.93,2.17] |
| Other living arrangements | | | | 0.86 [0.49,1.51] |
| Once infrequent contacts | | | | 0.83 [0.58,1.18] |
| Infrequent contacts throughout pandemic | | | | 0.73 [0.47,1.14] |
| Once high loneliness | | | | 4.68*** [3.68,5.94] |
| High loneliness throughout pandemic | | | | 14.29*** [11.03,18.52] |
| *Observations* | *5145* | *5006* | *4922* | *4862* |

Sources: ELSA, COVID-19 sub-study Wave 2 (November/December 2020), COVID-19 sub-study Wave 1 (June/July 2020) and Wave 9 (2018/19). * $p < 0.05$, ** $p < 0.01$, *** $p < 0.001$. Weighted data.

***Supplementary Table S4. Associations between shielding patterns and anxiety. Nested fully-adjusted logistic regression models – Odds Ratios [and 95% CIs]***

| | **Model 1** | **Model 2** | **Model 3** | **Model 4** |
|---|---|---|---|---|
| Staying at home at all three points | 1.84*** [1.33,2.56] | 1.49* [1.03,2.17] | 1.24 [0.85,1.81] | 1.18 [0.80,1.74] |
| Never shielding nor staying home at all points | Ref | Ref | Ref | Ref |
| Shielding in one time point | 3.82*** [2.48,5.88] | 2.23*** [1.44,3.45] | 1.74* [1.10,2.74] | 1.63* [1.00,2.64] |
| Shielding in two time points | 4.35*** [2.72,6.94] | 2.26** [1.35,3.77] | 2.22** [1.29,3.82] | 2.06* [1.16,3.65] |
| Shielding at all three time points | 4.37*** [2.54,7.52] | 1.71 [0.99,2.94] | 1.38 [0.77,2.47] | 1.46 [0.82,2.61] |
| Female | 1.80*** [1.36,2.39] | 1.82*** [1.34,2.46] | 1.79*** [1.30,2.46] | 1.61** [1.14,2.27] |
| Age | 0.95*** [0.94,0.97] | 0.95*** [0.93,0.97] | 0.95*** [0.93,0.97] | 0.96** [0.94,0.99] |
| Age squared | 1.00 [1.00,1.00] | 1.00 [1.00,1.00] | 1.00 [1.00,1.00] | 1.00 [1.00,1.00] |
| Non-White | 1.83* [1.07,3.13] | 1.35 [0.78,2.36] | 1.38 [0.77,2.47] | 1.16 [0.62,2.19] |
| Medium education | | 1.19 [0.82,1.73] | 1.33 [0.92,1.94] | 1.31 [0.89,1.92] |
| Low education | | 1.65* [1.12,2.45] | 1.82** [1.21,2.74] | 2.13*** [1.39,3.26] |
| 2nd lowest quintile | | 0.73 [0.47,1.11] | 0.80 [0.50,1.26] | 0.81 [0.51,1.30] |
| 3rd wealth quintile | | 0.76 [0.50,1.15] | 0.90 [0.58,1.40] | 0.95 [0.58,1.54] |
| 4th wealth quintile | | 0.75 [0.47,1.21] | 0.82 [0.50,1.36] | 0.91 [0.55,1.52] |
| Highest wealth quintile | | 0.84 [0.49,1.45] | 0.93 [0.53,1.64] | 0.99 [0.55,1.80] |
| Income | | 0.87 [0.73,1.04] | 0.89 [0.72,1.09] | 0.91 [0.75,1.11] |
| Employed not Working From Home (WFH) | | 0.83 [0.49,1.39] | 0.69 [0.40,1.19] | 0.72 [0.40,1.30] |
| Employed mostly WFH | | 0.72 [0.39,1.32] | 0.72 [0.38,1.36] | 0.75 [0.39,1.44] |
| Furloughed | | 0.48 [0.19,1.17] | 0.67 [0.28,1.65] | 0.75 [0.29,1.92] |
| Other employment | | 2.12** [1.32,3.40] | 2.09** [1.26,3.47] | 2.09** [1.24,3.54] |
| Homeowner with mortgage | | 0.84 [0.54,1.32] | 0.73 [0.45,1.17] | 0.74 [0.44,1.22] |
| Rented accommodation | | 0.92 [0.63,1.34] | 0.89 [0.59,1.34] | 0.95 [0.62,1.45] |
| Clinically vulnerable to COVID-19 | | 1.51** [1.14,2.02] | 1.46* [1.09,1.94] | 1.35 [0.99,1.85] |
| Disability | | 2.17*** [1.60,2.94] | 2.17*** [1.58,2.98] | 1.81*** [1.30,2.51] |
| Pre-pandemic Anxiety | | | 3.40*** [2.26,5.11] | 3.13*** [2.03,4.81] |
| Living Alone | | | | 0.63* [0.45,0.90] |
| Living with partner and children | | | | 1.32 [0.85,2.07] |
| Single parent | | | | 0.99 [0.50,1.95] |
| Other living arrangements | | | | 0.89 [0.37,2.13] |
| Once infrequent contacts | | | | 0.73 [0.40,1.33] |
| Infrequent contacts throughout pandemic | | | | 0.69 [0.34,1.38] |
| Once high loneliness | | | | 4.04*** [2.80,5.83] |
| High loneliness throughout pandemic | | | | 6.40*** [4.55,9.01] |
| *Observations* | *5142* | *4926* | *4596* | *4540* |

Sources: ELSA, COVID-19 sub-study Wave 2 (November/December 2020), COVID-19 sub-study Wave 1 (June/July 2020) and Wave 9 (2018/19). * $p < 0.05$, ** $p < 0.01$, *** $p < 0.001$. Weighted data

### *Supplementary Table S5. Associations between shielding patterns and quality of life. Nested fully-adjusted linear regression models – Beta coefficients [and 95% CIs]*

| | **Model 1** | **Model 2** | **Model 3** | **Model 4** |
|---|---|---|---|---|
| Staying at home at all three points | -1.048*** [-1.62,-0.47] | -0.407 [-0.94,0.13] | -0.468* [-0.87,-0.07] | -0.372* [-0.74,-0.00] |
| Never shielding nor staying home at all points | Ref | Ref | Ref | Ref |
| Shielding in one time point | -3.064*** [-3.94,-2.19] | -1.288*** [-2.03,-0.55] | -0.692* [-1.29,-0.09] | -0.531 [-1.11,0.05] |
| Shielding in two time points | -3.456*** [-4.29,-2.62] | -1.356*** [-2.11,-0.60] | -0.570 [-1.20,0.06] | -0.532 [-1.15,0.09] |
| Shielding at all three time points | -5.694*** [-7.06,-4.33] | -2.399*** [-3.60,-1.20] | -0.970* [-1.89,-0.05] | -1.283*** [-2.04,-0.52] |
| Female | -0.678** [-1.16,-0.19] | -0.569* [-1.01,-0.13] | -0.704*** [-1.04,-0.37] | -0.367* [-0.68,-0.06] |
| Age | 0.0263 [-0.00,0.05] | 0.0152 [-0.02,0.05] | 0.0368** [0.01,0.06] | 0.006 [-0.02,0.03] |
| Age squared | -0.006*** [-0.01,-0.00] | -0.004*** [-0.01,-0.00] | -0.002* [-0.00,-0.00] | -0.002** [-0.00,-0.00] |
| Non-White | -0.337 [-1.68,1.01] | 1.031 [-0.05,2.11] | 1.088* [0.01,2.16] | 1.375* [0.28,2.47] |
| Medium education | | 0.0254 [-0.51,0.56] | 0.049 [-0.35,0.44] | 0.049 [-0.31,0.41] |
| Low education | | 0.776* [0.13,1.42] | 0.868** [0.34,1.39] | 0.602* [0.11,1.09] |
| 2nd lowest quintile | | 1.402** [0.56,2.24] | 0.383 [-0.27,1.04] | 0.500 [-0.13,1.13] |
| 3rd wealth quintile | | 1.690*** [0.86,2.52] | 0.329 [-0.32,0.98] | 0.266 [-0.36,0.89] |
| 4th wealth quintile | | 1.938*** [1.10,2.78] | 0.202 [-0.46,0.86] | 0.193 [-0.43,0.81] |
| Highest wealth quintile | | 2.479*** [1.63,3.33] | 0.560 [-0.13,1.25] | 0.616 [-0.03,1.27] |
| Income | | 0.222*** [0.11,0.33] | 0.0256 [-0.04,0.09] | 0.021 [-0.04,0.08] |
| Employed not Working From Home (WFH) | | 0.254 [-0.55,1.06] | 0.478 [-0.10,1.05] | 0.272 [-0.27,0.81] |
| Employed mostly WFH | | 0.761 [-0.08,1.60] | 0.749* [0.11,1.39] | 0.673* [0.07,1.27] |
| Furloughed | | -1.599* [-2.99,-0.21] | -1.674** [-2.94,-0.41] | -1.793** [-2.98,-0.60] |
| Other employment | | -2.784*** [-3.96,-1.61] | -0.126 [-0.98,0.73] | -0.212 [-1.00,0.58] |
| Homeowner with mortgage | | -0.244 [-0.95,0.46] | -0.047 [-0.61,0.51] | -0.050 [-0.57,0.47] |
| Rented accommodation | | -0.512 [-1.29,0.26] | 0.032 [-0.59,0.65] | -0.098 [-0.71,0.51] |
| Clinically vulnerable to COVID-19 | | -1.311*** [-1.81,-0.81] | -0.237 [-0.61,0.14] | -0.185 [-0.52,0.15] |
| Disability | | -4.046*** [-4.64,-3.45] | -1.184*** [-1.67,-0.70] | -1.144*** [-1.60,-0.69] |
| Pre-pandemic Quality of Life | | | 0.687*** [0.65,0.72] | 0.564*** [0.53,0.60] |
| Living Alone | | | | 0.629*** [0.26,1.00] |
| Living with partner and children | | | | -0.436 [-0.95,0.08] |
| Single parent | | | | 0.054 [-0.80,0.91] |
| Other living arrangements | | | | 0.449 [-0.51,1.41] |
| Once infrequent contacts | | | | -0.545 [-1.18,0.09] |
| Infrequent contacts throughout pandemic | | | | 0.302 [-0.54,1.15] |
| Once high loneliness | | | | -2.803*** [-3.30,-2.31] |
| High loneliness throughout pandemic | | | | -4.953*** [-5.48,-4.42] |
| Constant | 27.51*** [27.01,28.01] | 25.66*** [24.60,26.71] | 7.665*** [6.44,8.89] | 11.850*** [10.56,13.13] |
| *Observations* | *5146* | *5007* | *4676* | *4619* |

Sources: ELSA, COVID-19 sub-study Wave 2 (November/December 2020), COVID-19 sub-study Wave 1 (June/July 2020) and Wave 9 (2018/19). * $p < 0.05$, ** $p < 0.01$, *** $p < 0.001$. Weighted data

***Supplementary Table S6. Associations between shielding patterns and life satisfaction. Nested fully-adjusted linear regression models – Beta coefficients [and 95% CIs]***

| | **Model 1** | **Model 2** | **Model 3** | **Model 4** |
|---|---|---|---|---|
| Staying at home at all three points | -0.282** [-0.48,-0.08] | -0.116 [-0.31,0.08] | -0.147 [-0.32,0.02] | -0.114 [-0.28,0.05] |
| Never shielding nor staying home at all points | Ref | Ref | Ref | Ref |
| Shielding in one time point | -0.508*** [-0.79,-0.22] | -0.184 [-0.44,0.07] | -0.080 [-0.32,0.16] | -0.007 [-0.24,0.22] |
| Shielding in two time points | -0.505*** [-0.79,-0.22] | -0.113 [-0.40,0.17] | -0.145 [-0.40,0.12] | -0.068 [-0.33,0.20] |
| Shielding at all three time points | -0.918*** [-1.42,-0.42] | -0.359 [-0.83,0.11] | -0.282 [-0.68,0.12] | -0.273 [-0.65,0.10] |
| Female | -0.208* [-0.38,-0.04] | -0.221** [-0.39,-0.05] | -0.193* [-0.34,-0.04] | -0.080 [-0.23,0.07] |
| Age | 0.029*** [0.02,0.04] | 0.028*** [0.01,0.04] | 0.020*** [0.01,0.03] | 0.014** [0.00,0.02] |
| Age squared | -0.001 [-0.00,0.00] | -0.000 [-0.00,0.00] | -0.000 [-0.00,0.00] | -0.000 [-0.00,0.00] |
| Non-White | -0.189 [-0.65,0.28] | 0.020 [-0.40,0.45] | 0.092 [-0.31,0.50] | 0.195 [-0.26,0.65] |
| Medium education | | 0.148 [-0.04,0.33] | 0.042 [-0.13,0.21] | 0.064 [-0.10,0.23] |
| Low education | | 0.558*** [0.33,0.78] | 0.412*** [0.21,0.61] | 0.364*** [0.17,0.56] |
| 2nd lowest quintile | | 0.212 [-0.11,0.53] | 0.011 [-0.29,0.31] | -0.023 [-0.33,0.28] |
| 3rd wealth quintile | | 0.087 [-0.22,0.39] | -0.117 [-0.39,0.15] | -0.215 [-0.49,0.05] |
| 4th wealth quintile | | 0.165 [-0.15,0.48] | -0.063 [-0.35,0.22] | -0.169 [-0.45,0.11] |
| Highest wealth quintile | | 0.215 [-0.09,0.52] | -0.125 [-0.40,0.15] | -0.200 [-0.48,0.08] |
| Income | | 0.052** [0.01,0.09] | 0.023 [-0.01,0.05] | 0.016 [-0.01,0.04] |
| Employed not Working From Home (WFH) | | 0.129 [-0.18,0.44] | 0.150 [-0.11,0.41] | 0.085 [-0.17,0.34] |
| Employed mostly WFH | | 0.225 [-0.09,0.54] | 0.202 [-0.07,0.48] | 0.178 [-0.09,0.44] |
| Furloughed | | 0.075 [-0.53,0.68] | 0.257 [-0.21,0.72] | 0.202 [-0.23,0.63] |
| Other employment | | -0.618** [-1.04,-0.20] | -0.186 [-0.54,0.17] | -0.154 [-0.51,0.20] |
| Homeowner with mortgage | | 0.044 [-0.24,0.33] | -0.021 [-0.28,0.23] | -0.024 [-0.28,0.23] |
| Rented accommodation | | -0.192 [-0.49,0.11] | -0.090 [-0.36,0.18] | -0.082 [-0.35,0.19] |
| Clinically vulnerable to COVID-19 | | -0.146 [-0.32,0.03] | 0.038 [-0.12,0.20] | 0.082 [-0.08,0.24] |
| Disability | | -0.832*** [-1.06,-0.61] | -0.363*** [-0.56,-0.16] | -0.311** [-0.51,-0.12] |
| Pre-pandemic Quality of Life | | | 0.438*** [0.40,0.48] | 0.335*** [0.29,0.38] |
| Living Alone | | | | 0.030 [-0.15,0.21] |
| Living with partner and children | | | | -0.143 [-0.38,0.10] |
| Single parent | | | | -0.161 [-0.51,0.19] |
| Other living arrangements | | | | 0.184 [-0.21,0.58] |
| Once infrequent contacts | | | | -0.125 [-0.40,0.15] |
| Infrequent contacts throughout pandemic | | | | -0.037 [-0.37,0.30] |
| Once high loneliness | | | | -0.865*** [-1.07,-0.66] |
| High loneliness throughout pandemic | | | | -1.602*** [-1.83,-1.37] |
| Constant | 7.387*** [7.21,7.56] | 6.964*** [6.60,7.32] | 3.811*** [3.38,4.24] | 4.935*** [4.49,5.39] |
| Observations | 5136 | 4997 | 4624 | 4567 |

Sources: ELSA, COVID-19 sub-study Wave 2 (November/December 2020), COVID-19 sub-study Wave 1 (June/July 2020) and Wave 9 (2018/19)
